# Can Artificial Intelligence Match Dermoscopy in Melanoma Detection? Evidence from a Systematic Review and Meta-analysis of Pigmented Skin Lesions

**DOI:** 10.64898/2026.05.15.26353363

**Authors:** Hailing Tang, Yaxi Zhu, Ming Diao

## Abstract

**Background:** Accurate risk stratification of pigmented skin lesions is essential for early melanoma detection and reducing unnecessary excisions. Although artificial intelligence (AI) is increasingly applied to dermoscopic image analysis, its diagnostic performance relative to dermoscopy remains uncertain.

**Objective:** To compare the diagnostic performance of AI, dermoscopy, and AI-assisted clinicians in the malignancy risk stratification of pigmented skin lesions.

**Methods:** PubMed, Embase, Web of Science, and Cochrane Library were systematically searched for studies evaluating AI, dermoscopy, or AI-assisted clinicians in diagnosing pigmented or melanoma-suspected skin lesions. Diagnostic performance metrics were calculated from extracted or reconstructed data, and study quality was assessed using QUADAS-2 and QUADAS-C.

**Results:** A total of 2571 records were identified, and 10 studies were included in the main quantitative analysis, contributing 17 diagnostic arms. These included 10 dermoscopy arms, 6 AI-alone arms, and 1 AI-assisted clinician arm. In the dermoscopy group, sensitivity ranged from 0.418 to 0.966, and specificity ranged from 0.293 to 0.975. In the AI group, sensitivity ranged from 0.164 to 0.968, and specificity ranged from 0.374 to 0.983. AI-assisted clinicians showed a sensitivity of 1.000 and specificity of 0.837 in the single available study. Overall, AI and dermoscopy showed overlapping diagnostic performance, although substantial variability was observed across AI algorithms and clinical settings. Deeks funnel plots did not indicate significant publication bias in either the AI group or the dermoscopy group.

**Conclusions:** Autonomous AI showed diagnostic performance broadly comparable to dermoscopy. Although AI achieved slightly higher pooled specificity, its sensitivity was lower, indicating no clear clinical advantage. AI is therefore best used as an adjunct to dermatological evaluation rather than a standalone tool.

## 1. Introduction

Malignant melanoma is among the most aggressive forms of skin cancer, making early detection and timely surgical intervention crucial for improving patient outcomes. In routine dermatologic practice, pigmented skin lesions encompass a highly heterogeneous diagnostic spectrum, seamlessly bridging innocuous melanocytic nev[1], dysplastic proliferations, and potentially fatal melanomas[2][3]. Consequently, accurate malignancy risk stratification represents a central clinical imperative. Practitioners face the dual challenge of maximizing diagnostic sensitivity to avoid the catastrophic consequences of missed melanomas, while simultaneously maintaining sufficient specificity to reduce the physical morbidity and economic burden associated with the unnecessary excision of benign mimickers [4][5].

To navigate this morphological complexity, dermoscopy, a noninvasive diagnostic technique, has become an indispensable tool in dermatology by enabling improved visualization of subsurface skin structures. Compared to unaided visual inspection, it substantially increases diagnostic accuracy for melanoma and helps to reduce unnecessary excisions of benign lesions [6][7]. Despite being the current standard of care, however, its diagnostic performance remains highly dependent on the clinician’s level of experience. Pattern recognition and heuristic evaluations vary significantly; thus, the diagnostic yield achieved by an expert dermoscopist frequently eclipses that of a general practitioner or a dermatologist with limited optical training [8][9].

In recent years, artificial intelligence (AI) systems have shown promising results in the automated analysis and classification of dermoscopic images. Convolutional neural networks (CNNs), in particular, have become the most commonly used approach in this field and form the basis for most modern algorithms for classifying skin lesions. Numerous retrospective studies have reported diagnostic performances comparable to, or even exceeding, those of expert dermatologists [10][11]. Nevertheless, most studies evaluating AI performance have been retrospective and rely on curated image datasets that may not reflect the complexity of everyday clinical practice. These retrospective analyses are limited in their ability to assess generalizability, risk of bias, and true diagnostic impact, frequently overestimating the algorithm’s translational accuracy when confronted with diverse skin-tone populations or unrepresented lesion morphologies[12][13].

Historically, the discourse surrounding machine learning in dermatology has fixated on an adversarial narrative, primarily focusing on direct comparisons between isolated AI algorithms and clinicians [14]. Considerably less attention has been paid to the pragmatic integration of AI as a synergistic overlay—specifically, AI-assisted clinicians—designed to augment human decision-making, or as a benchmarked alternative to standalone dermoscopy, a mature noninvasive diagnostic technique [15]. While individual studies have shown promising results for AI in skin cancer, there remains a lack of systematic evidence directly comparing AI alone, standalone dermoscopy, and AI-assisted clinicians in prospective clinical settings.[16]. By synthesizing available evidence, this systematic review and meta-analysis aims to compare the diagnostic performance of AI alone, standalone dermoscopy, and AI-assisted clinicians for the malignancy risk stratification of pigmented skin lesions, ultimately informing the current state of clinical readiness for these technologies in real-world settings.

## 2. Methods

The systematic review and meta-analysis adhered to the guidelines outlined in the Preferred Reporting Items for Systematic Reviews and Meta-Analyses (PRISMA) statement. The study was registered in advance on PROSPERO (CRD420261389834).

### 2.1 Eligibility criteria

We included diagnostic accuracy studies that evaluated AI, dermoscopy, or AI-assisted clinicians for malignancy risk stratification of pigmented skin lesions. Eligible studies enrolled patients with pigmented, melanocytic, or melanoma-suspected skin lesions and reported diagnostic performance data for at least one of the following diagnostic approaches: AI alone, dermoscopy or dermatologist dermoscopic assessment, or dermatologists assisted by AI. Studies were eligible if the reference standard was histopathological examination, or histopathology combined with clinical follow-up or expert consensus for nonexcised benign lesions. Studies were required to provide sufficient quantitative data to extract or calculate true positives (TP), false positives (FP), false negatives (FN), and true negatives (TN), or to report sensitivity and specificity that could be converted into a 2 × 2 diagnostic table.

The primary analysis focused on prospective clinical studies or prospective diagnostic validation studies using dermoscopic images or dermoscopy-based assessment. Diagnostic arms were categorized into three groups: AI alone, dermoscopy, and dermatologist assisted by AI. Studies evaluating nondermoscopic AI applications, mobile applications, reader studies, or non-AI imaging techniques were considered for supplementary analysis when relevant, but were not combined with the main pooled analysis.

We excluded studies based on the following criteria: (1) reviews, editorials, commentaries, letters, conference abstracts, or protocols; (2) purely algorithm-development studies without clinical validation; (3) studies using only public retrospective image datasets without a clinical diagnostic setting; (4) studies not involving pigmented skin lesions, melanocytic lesions, melanoma, or skin cancer risk stratification; (5) studies without an appropriate reference standard; (6) studies with insufficient data to extract or calculate diagnostic accuracy outcomes; or (7) duplicated study populations, in which case the most complete or most clinically relevant report was retained.

### 2.2 Search strategy and study selection

A systematic literature search was conducted in PubMed, Embase, Web of Science, and the Cochrane Library from database inception to January 2026. The search strategy combined controlled vocabulary terms and free-text terms related to artificial intelligence, dermoscopy, pigmented skin lesions, melanoma, and diagnostic accuracy. The main search terms included “artificial intelligence,” “machine learning,” “deep learning,” “convolutional neural network,” “dermoscopy,” “dermatoscopy,” “pigmented skin lesion,” “melanocytic lesion,” “melanoma,” “skin cancer,” “diagnostic accuracy,” and “prospective study.” Boolean operators were used to combine search terms.

All retrieved records were imported into EndNote™ (Version X7) for deduplication and reference management [17]. Study selection was conducted in two stages. First, titles and abstracts were screened to exclude clearly irrelevant publications. Second, full-text articles were reviewed according to the predefined inclusion and exclusion criteria. Disagreements were resolved through discussion, and uncertain studies were rechecked before final inclusion. The study selection process was summarized using a PRISMA flow diagram.

### 2.3 Data extraction and quality assessment

Data were extracted using a standardized data extraction form. The extracted information included the first author, publication year, country or region, study design, sample size, number of patients and lesions, number of melanoma or malignant lesions, number of benign or nonmelanoma lesions, diagnostic approach, AI system or dermoscopy method, comparator, reference standard, and follow-up method when applicable. For each eligible diagnostic arm, TP, FP, FN, and TN were extracted directly when available. When a 2 × 2 table was not explicitly reported, the values were recalculated from reported sensitivity, specificity, and the number of malignant and nonmalignant lesions.

Diagnostic arms were classified into three predefined groups: AI alone, dermoscopy, and dermatologist assisted by AI. For studies reporting multiple AI algorithms or multiple clinician groups, each diagnostic arm was extracted separately. However, sensitivity analyses were planned to assess the influence of multiple arms derived from the same study population.

The methodological quality of included studies was assessed using the Quality Assessment of Diagnostic Accuracy Studies 2 tool (QUADAS-2) [18].Four domains were evaluated: patient selection, index test, reference standard, and flow and timing. Each domain was judged as low, high, or unclear risk of bias. Applicability concerns were assessed for patient selection, index test, and reference standard. For studies directly comparing AI with dermoscopy or dermatologist assessment within the same clinical setting, the QUADAS-C tool[19] was additionally used to evaluate the risk of bias in comparative diagnostic accuracy.

### 2.4 Statistical analysis

All statistical analyses were performed using R software (version 4.3.0; R Foundation for Statistical Computing, Vienna, Austria). For each diagnostic arm, true positives (TP), false positives (FP), false negatives (FN), and true negatives (TN) were extracted or reconstructed from the original studies. Sensitivity, specificity, accuracy, and balanced accuracy were calculated to evaluate diagnostic performance. Sensitivity was defined as TP/(TP + FN), specificity as TN/(TN + FP), accuracy as (TP + TN)/(TP + FP + FN + TN), and balanced accuracy as the average of sensitivity and specificity.

Diagnostic arms were categorized into three predefined groups: artificial intelligence (AI) alone, dermoscopy, and AI-assisted clinicians. Study-level sensitivity and specificity with 95% confidence intervals (CI) were visualized using forest plots. The distributions of sensitivity, specificity, accuracy, and balanced accuracy across diagnostic groups were compared using box plots. Summary receiver operating characteristic (SROC) curves were generated by plotting sensitivity against 1 − specificity to visually compare the overall diagnostic performance of AI and dermoscopy. Considering the heterogeneity in AI algorithms, dermoscopic devices, clinician experience, and reference standards across studies, pooled estimates were analyzed using a random-effects model. Potential publication bias and small-study effects were explored using Deeks’ funnel plots. All tests were two-sided, with P < 0.05 considered statistically significant[20].

## 3. Results

### 3.1 Study selection

A total of 2571 records were identified through database searching. After removing 657 duplicates, 1914 records were screened by title and abstract. Among them, 1806 records were excluded as irrelevant. Subsequently, 92 full-text articles were assessed for eligibility. Finally, 10 studies contributing 17 diagnostic arms were included in the main diagnostic performance analysis. The 17 diagnostic arms consisted of 10 dermoscopy arms, 6 AI-alone arms, and 1 AI-assisted clinician arm. The study selection process is shown in Figure 1.

**Figure 1.**
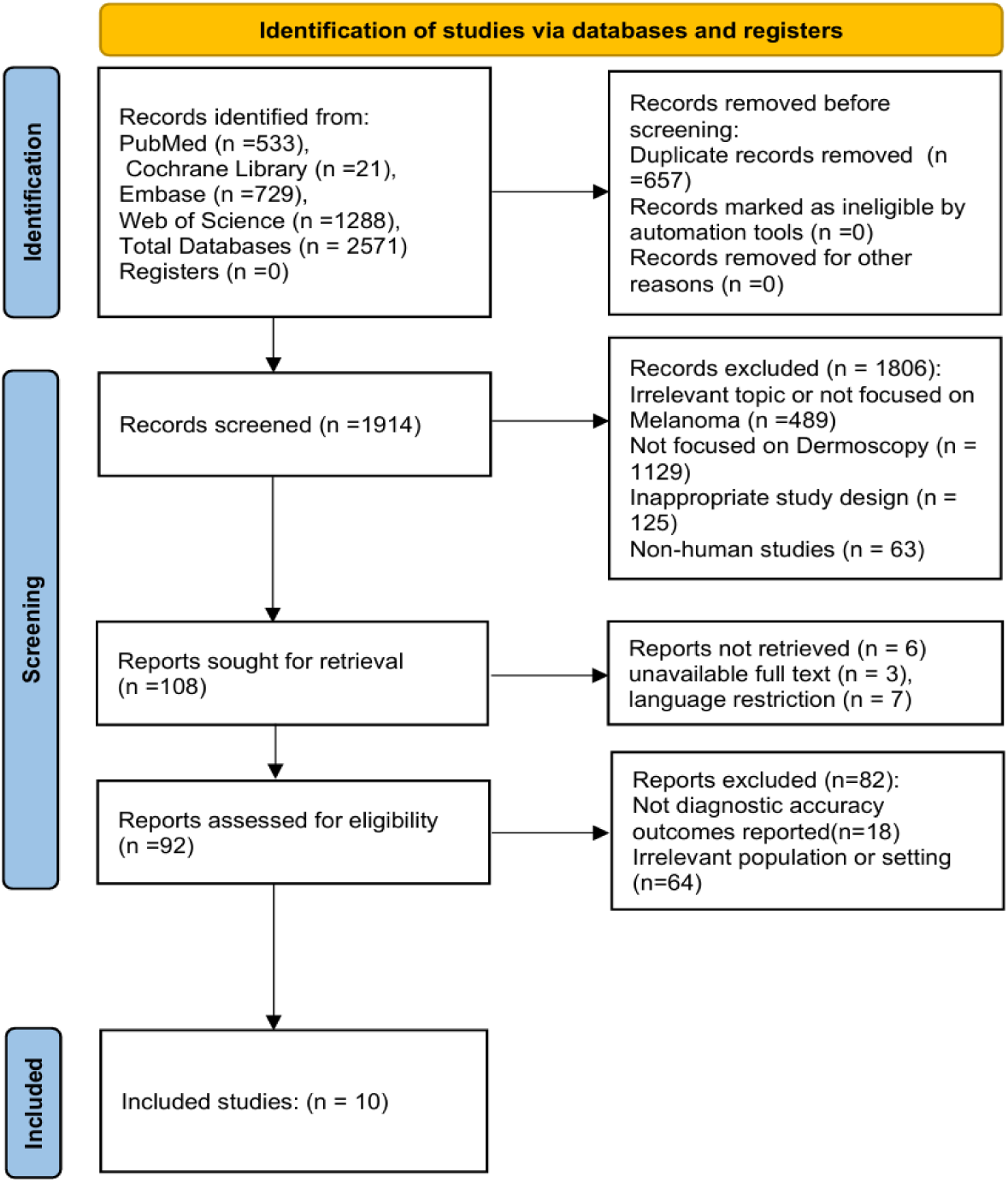
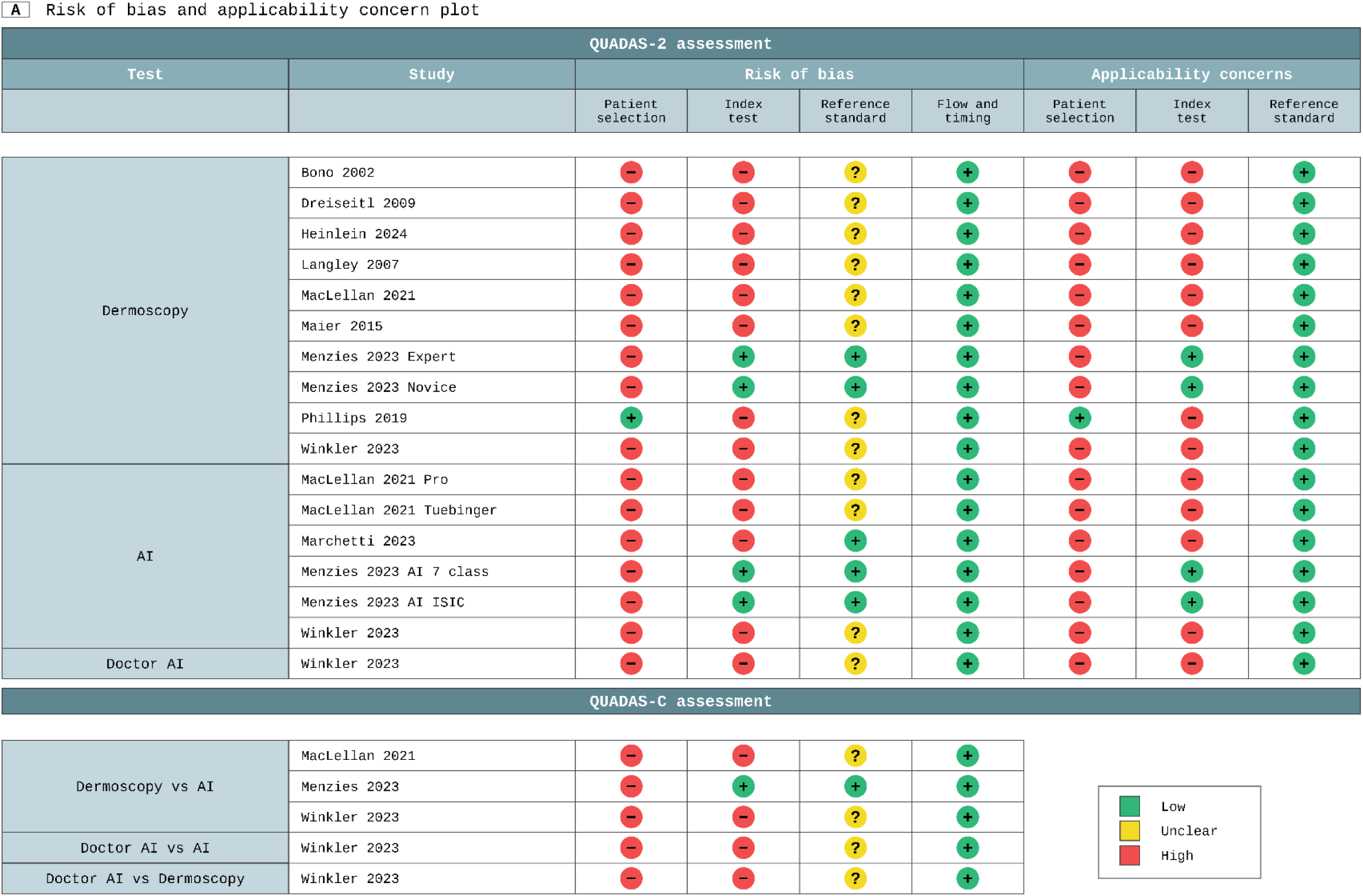
PRISMA flow diagram of study selection. A total of 2571 records were identified from PubMed, Cochrane Library, Embase, and Web of Science. After removing 657 duplicates, 1914 records were screened. Finally, 10 studies were included in the main quantitative analysis.

### 3.2 Study characteristics

A total of 10 studies published between 2002 and 2024 were included in the main quantitative analysis [16,21–29]. These studies were conducted in Italy, Austria, Germany, Canada, the United Kingdom, the United States, and Australia. Most were prospective diagnostic accuracy studies or prospective clinical validation studies, enrolling patients with clinically suspicious pigmented, melanocytic, or melanoma-suspected lesions.

Sample sizes varied across studies, with the number of lesions ranging from 125 to 1910. Histopathology was the main reference standard, while clinical follow-up or expert consensus was used for some nonexcised benign lesions. Diagnostic approaches were classified into three groups: dermoscopy, AI alone, and AI-assisted clinicians. The dermoscopy group included dermatologist or clinician assessment based on dermoscopy; the AI group included systems such as FotoFinder, ADAE, AI 7-class, AI ISIC, and Moleanalyzer Pro CNN; and only one study evaluated dermatologists assisted by AI. The characteristics of the included studies are shown in Table 1.

**Table 1.**
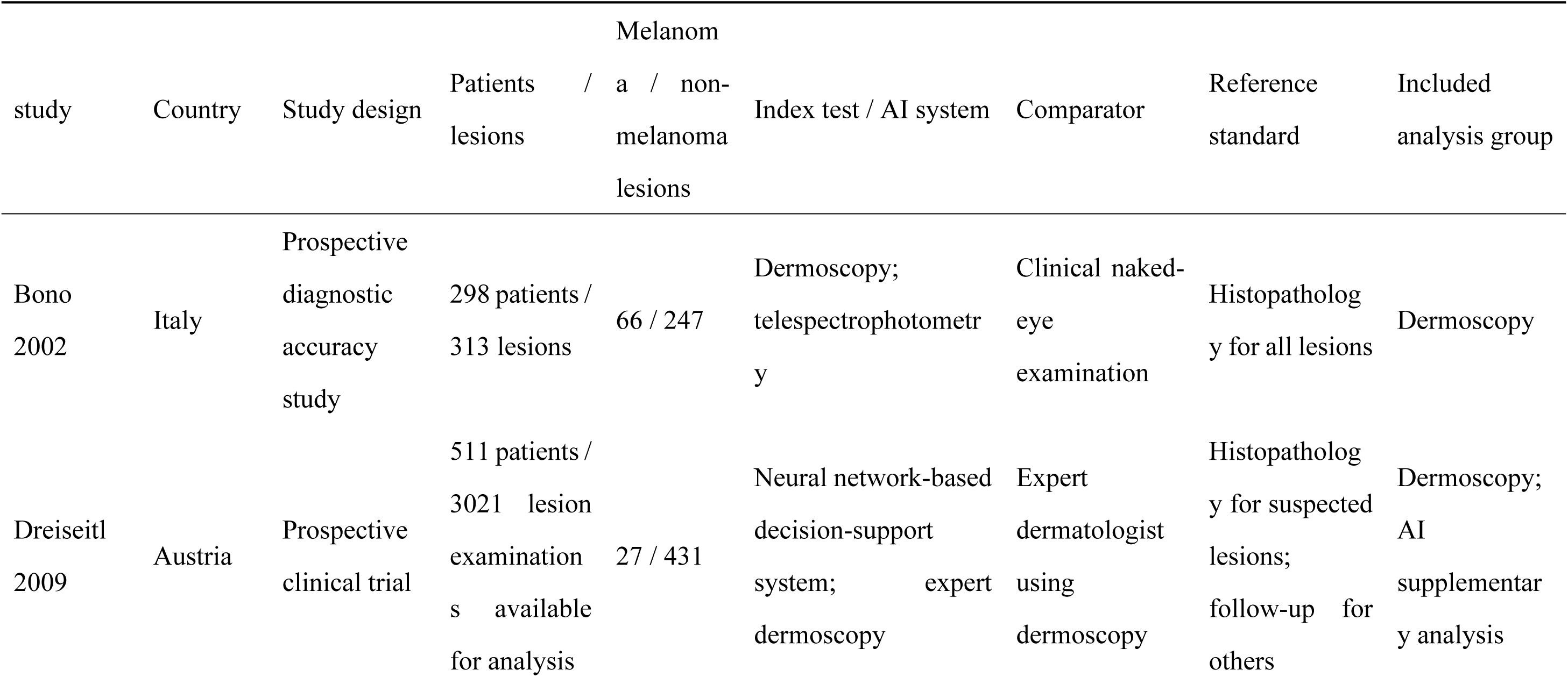
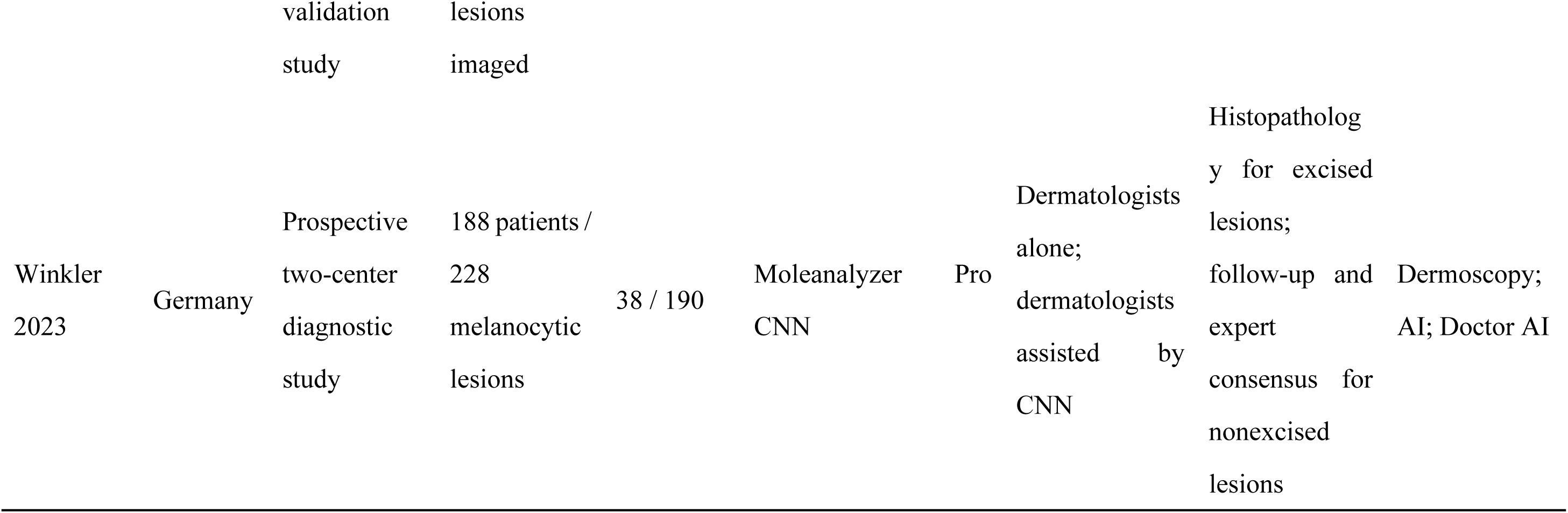
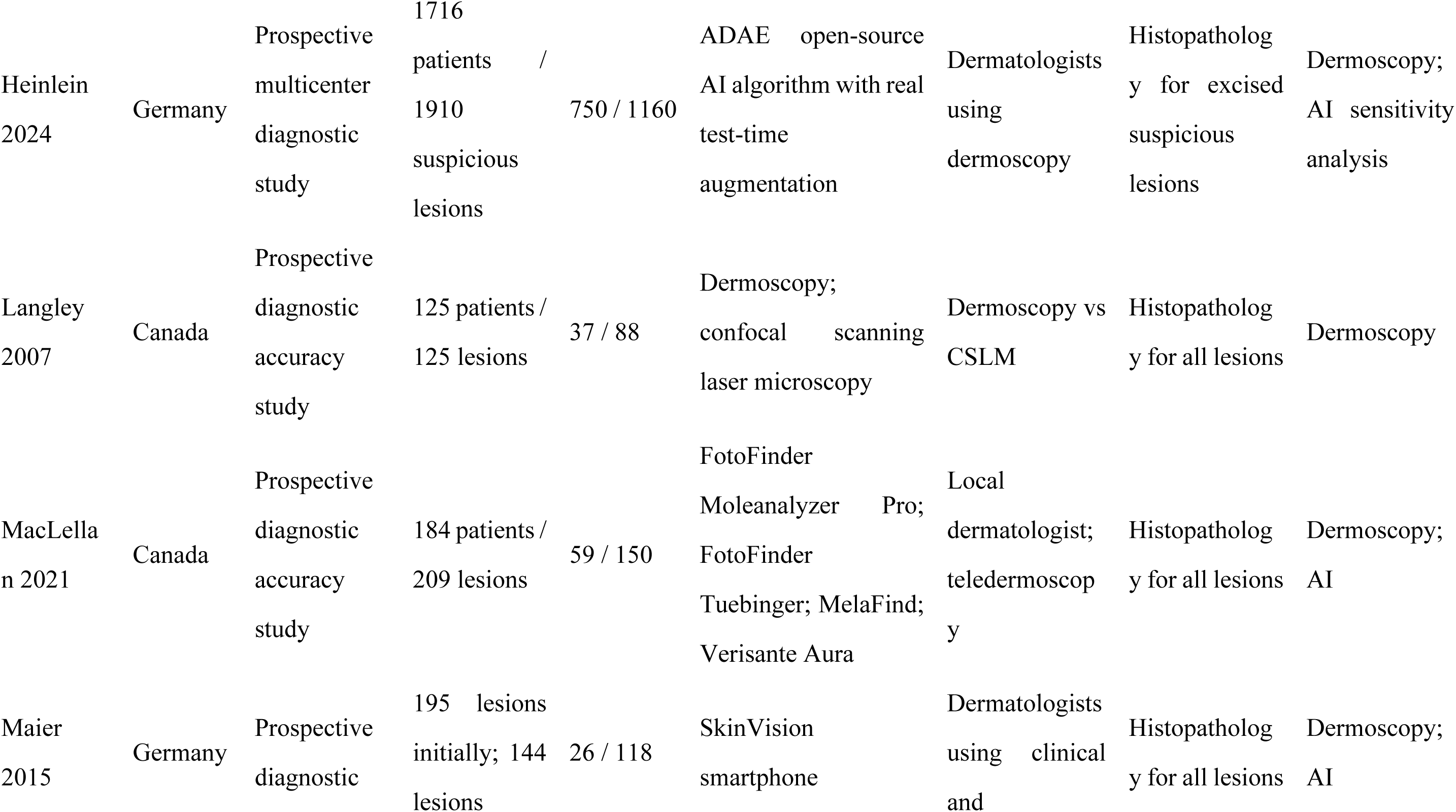
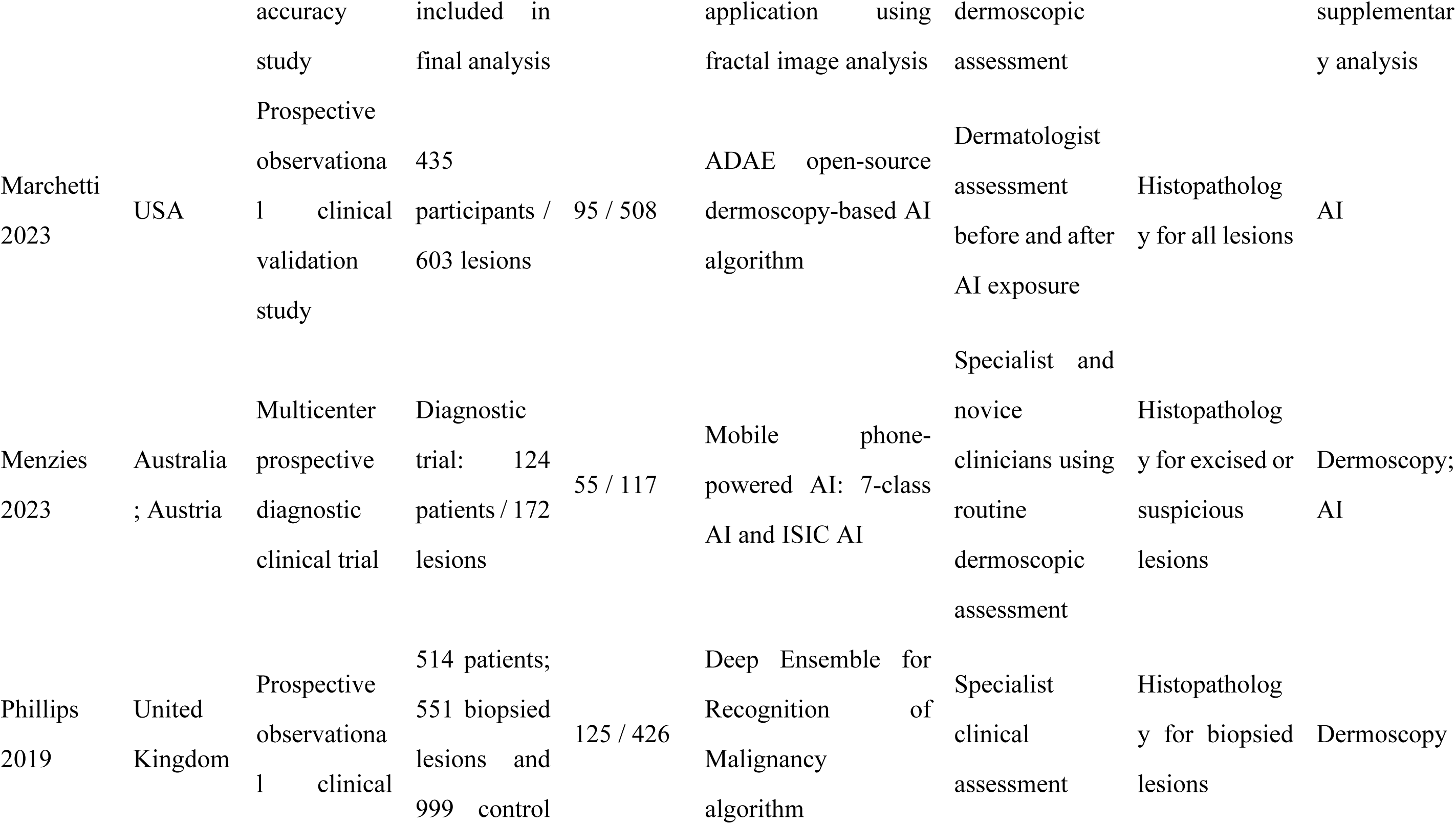
Characteristics of included studies. This table summarizes the main characteristics of the 10 studies included in the main quantitative analysis, including publication year, country or region, study design, sample size, diagnostic method, comparator, reference standard, and included analysis group.

### 3.3 Risk of bias and applicability concerns

QUADAS-2 and QUADAS-C were used to assess the methodological quality of the included studies. Overall, the main risk of bias came from patient selection. Most studies enrolled clinically suspicious or biopsy-planned pigmented lesions, rather than unselected screening populations. Therefore, patient selection was frequently judged as high risk. For the index test domain, several studies were rated as high risk because of differences in AI algorithms, diagnostic thresholds, clinician experience, and unclear blinding. The reference standard was generally acceptable because most studies used histopathology, although some were rated as unclear when blinding of pathological assessment was not reported. Flow and timing was mostly judged as low risk. Applicability concerns were mainly found in patient selection and index test domains, while concerns about the reference standard were generally low. The detailed QUADAS-2 and QUADAS-C results are shown in Figure 2.

**Figure 2.**
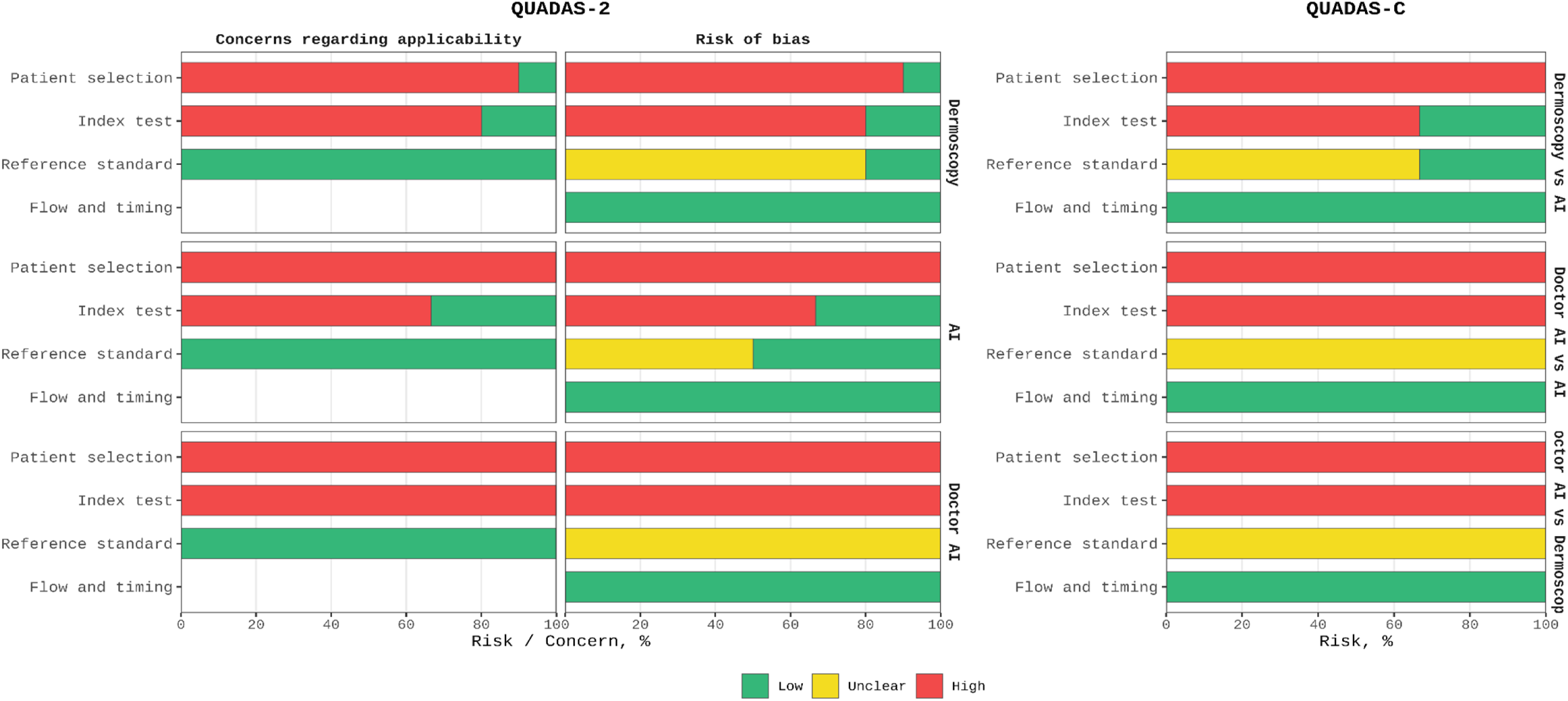
Risk of bias and applicability assessment of included studies. QUADAS-2 was used to assess risk of bias and applicability concerns across the included diagnostic arms. QUADAS-C was further applied to studies with direct comparisons between dermoscopy, AI, and AI-assisted clinicians. Green indicates low risk, yellow indicates unclear risk, and red indicates high risk.

### 3.4 Diagnostic performance and comparative meta-analysis

Figure 3 summarizes diagnostic accuracy across study arms. In the dermoscopy group (10 arms), a bivariate random-effects model yielded a pooled sensitivity of 0.773 (95% CI, 0.648–0.863, P < 0.001) and specificity of 0.793 (95% CI, 0.673–0.877, P < 0.001). Substantial between-study heterogeneity was observed, with sensitivity ranging from 0.418 to 0.966 and specificity from 0.293 to 0.975, suggesting variation associated with clinician experience and practice setting.

**Figure 3.**
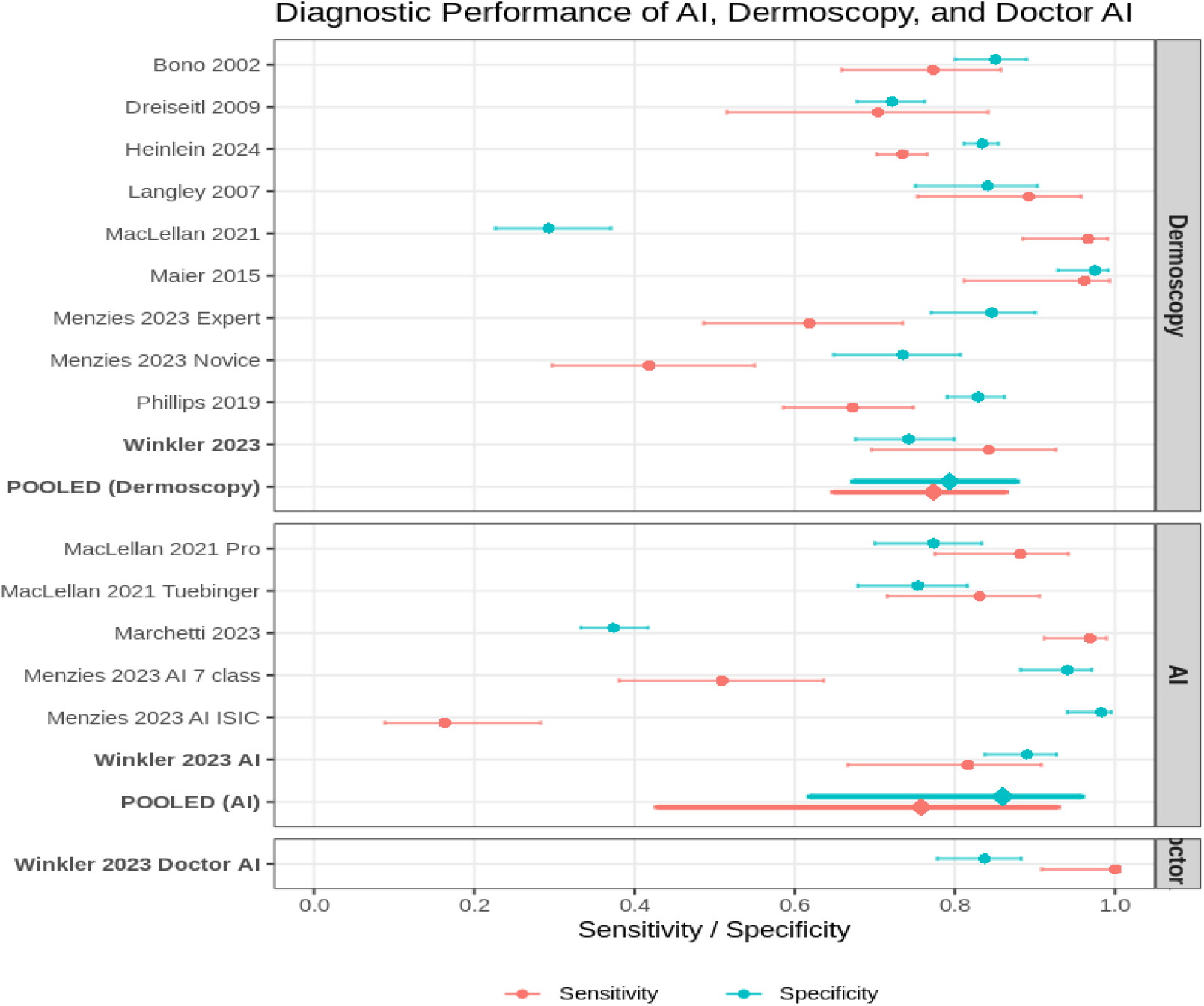
Study-level sensitivity and specificity of AI, dermoscopy, and AI-assisted clinicians. Each point represents the sensitivity or specificity of one diagnostic arm, with horizontal bars indicating 95% confidence intervals. Diagnostic arms were grouped as dermoscopy, AI alone, and AI-assisted clinicians.

In the standalone artificial intelligence (AI) group (6 arms), pooled sensitivity was 0.757 (95% CI, 0.428–0.928, P < 0.001) and specificity was 0.859 (95% CI, 0.619–0.958, P < 0.001). A threshold effect was evident across algorithms. The ADAE model favored sensitivity at the expense of specificity, whereas the AI-ISIC arm showed higher specificity with lower sensitivity, reflecting differences in model calibration. The performance of AI as an assistive tool was evaluated in Winkler (2023), in which AI-assisted dermatologists achieved a sensitivity of 1.000 and specificity of 0.837. Although this finding suggests a potential benefit over unaided assessment, the available evidence remains limited and requires prospective validation.

### 3.5 Heterogeneity and threshold effect analysis

Exploration of inter-study variance revealed substantial heterogeneity within both diagnostic modalities. Quantitative assessment using Holling’s unadjusted *I*² estimates demonstrated moderate to high heterogeneity, ranging from 54.3% to 87.9% in the dermoscopy cohort, and 74.7% to 81.2% in the standalone AI cohort. Given the inherent trade-off between sensitivity and specificity in diagnostic test accuracy studies, a bivariate correlation analysis was conducted to ascertain whether this variance was attributable to the threshold effect. The correlation coefficient between logit sensitivity and logit false-positive rate was 0.26 for dermoscopy, indicating that non-threshold factors (e.g., clinician experience, diverse patient demographics, and lesion complexity) primarily drove the observed variability. Conversely, the AI cohort exhibited a perfect positive correlation (*r* = 1.00). This salient finding indicates that the profound heterogeneity observed among AI algorithms is overwhelmingly driven by threshold effects. Specifically, the variance stems from discrepancies in the predefined operating cut-offs optimized by different developers, rather than fundamental disparities in the intrinsic discriminatory capacity of the neural networks themselves.

### 3.6 Diagnostic performance metrics

The box plot shows the distribution of accuracy, balanced accuracy, sensitivity, and specificity across the three diagnostic groups. Overall, AI and dermoscopy showed similar distributions in accuracy and balanced accuracy. However, the AI group showed wider variation in sensitivity and specificity, suggesting that diagnostic performance differed across AI systems. The dermoscopy group also showed variability, which may be related to differences in clinician experience, diagnostic thresholds, and study populations. The Doctor AI group showed high values across all four metrics, including sensitivity, specificity, accuracy, and balanced accuracy. However, this group included only one diagnostic arm, so the result should be interpreted cautiously. The distribution of diagnostic performance metrics is shown in Figure 4A.

**Figure 4.**
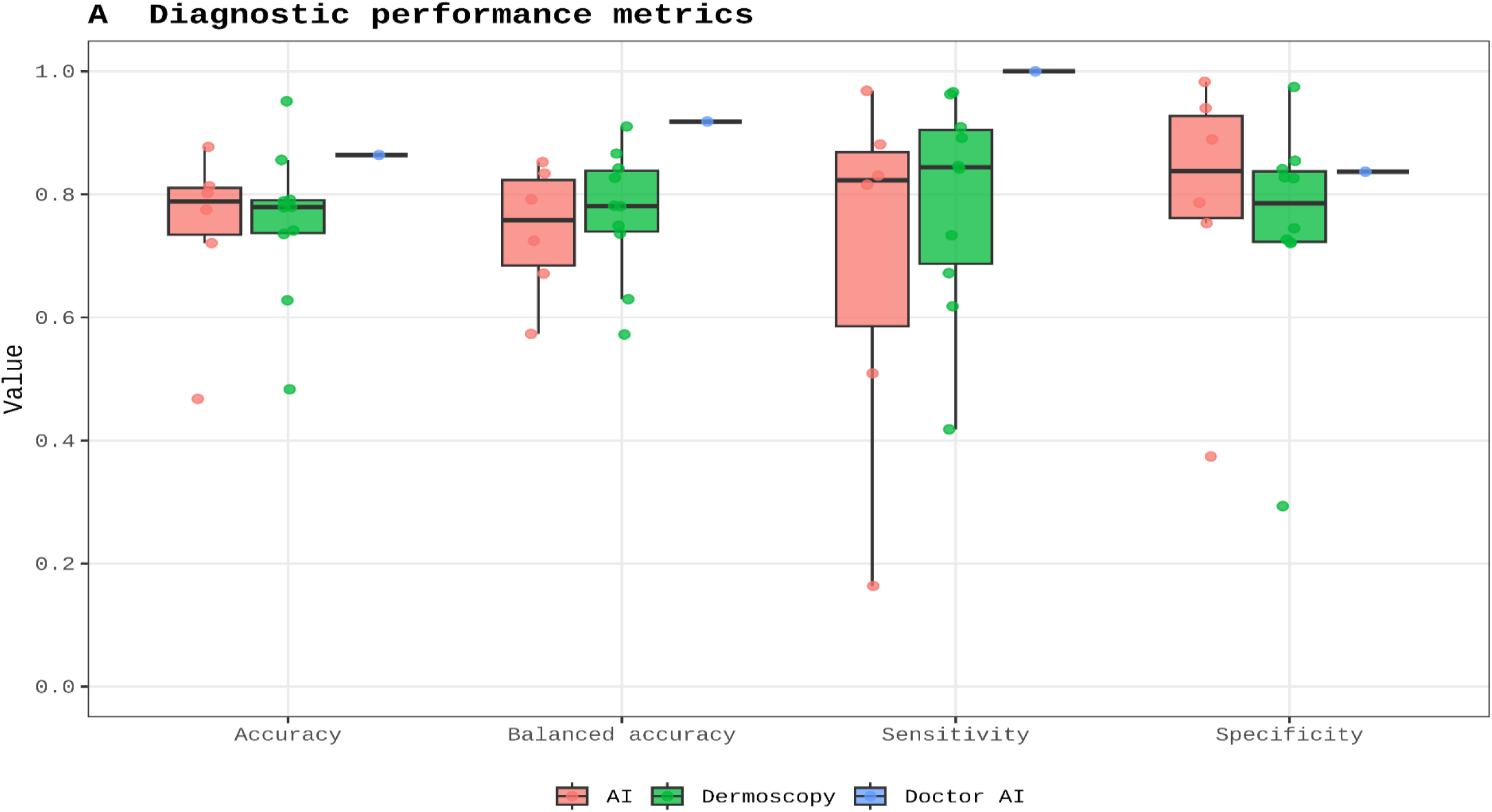
Distribution of diagnostic performance metrics. Box plots showing accuracy, balanced accuracy, sensitivity, and specificity across AI, dermoscopy, and AI-assisted clinician groups.

### 3.7 Summary ROC analysis

The SROC plot was used to visually compare the overall diagnostic performance of AI and dermoscopy. The study-level estimates of AI and dermoscopy were distributed in overlapping areas, and no clear separation between the two curves was observed. This suggests that AI alone had an overall diagnostic performance broadly comparable to dermoscopy. However, the distribution of AI points was relatively dispersed, indicating heterogeneity among different AI algorithms and diagnostic thresholds. Similarly, dermoscopy estimates also varied across studies, likely reflecting differences in clinician experience and study design. The SROC curves are shown in Figure 5.

**Figure 5.**
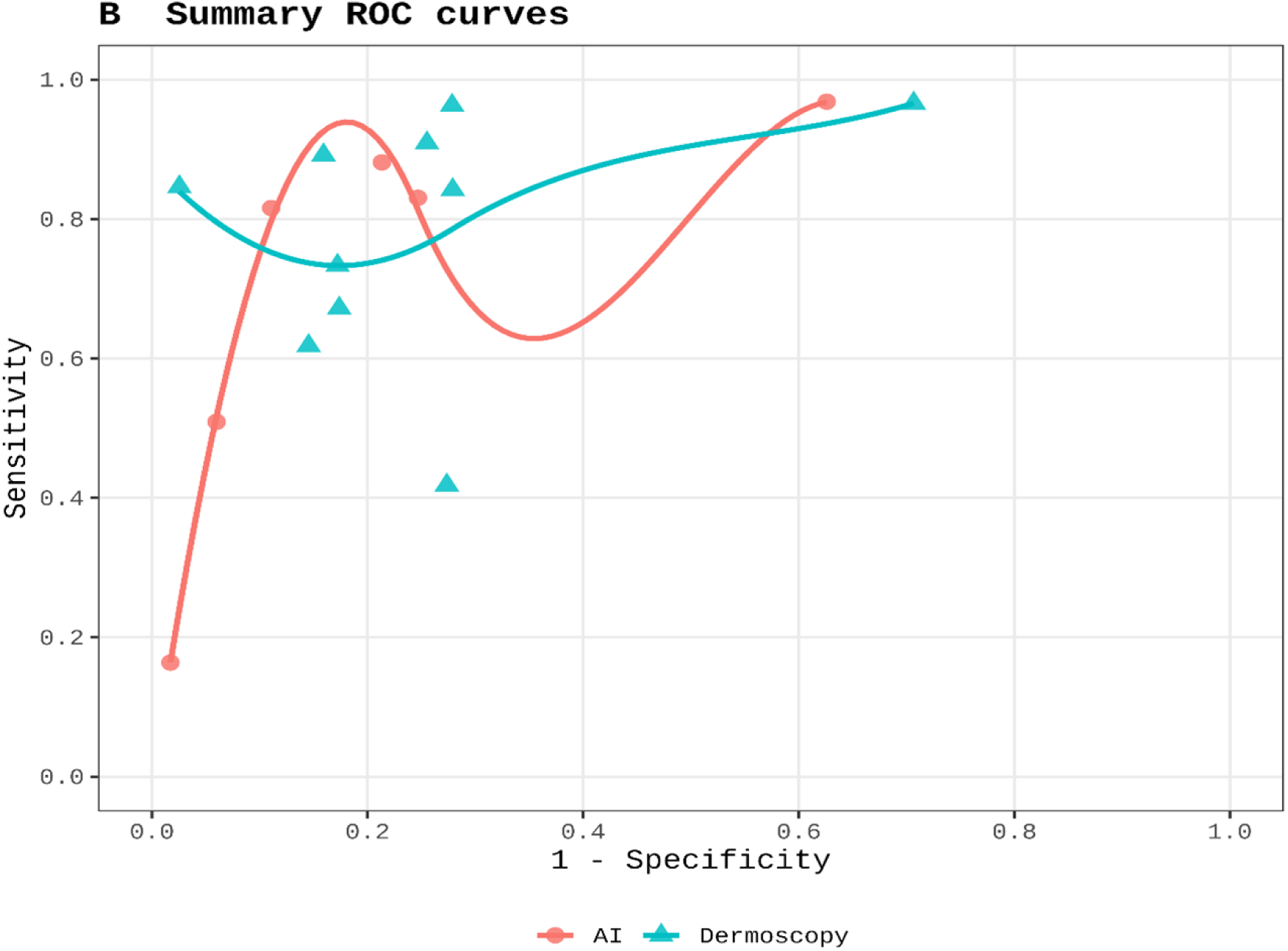
Summary ROC curves of AI and dermoscopy. The SROC plot compares study-level diagnostic performance between AI and dermoscopy using sensitivity and 1 − specificity.

### 3.8 Publication bias

Potential publication bias and small-study effects were assessed using Deeks’ funnel plots. In the AI group, the Deeks test showed no significant evidence of publication bias (P = 0.693). Similarly, no significant publication bias was detected in the dermoscopy group (P = 0.511). Visual inspection of the funnel plots also did not suggest obvious asymmetry. However, because the number of included diagnostic arms was limited, especially in the AI group, these results should be interpreted cautiously. The Deeks funnel plots are shown in Figure 6.

**Figure 6.**
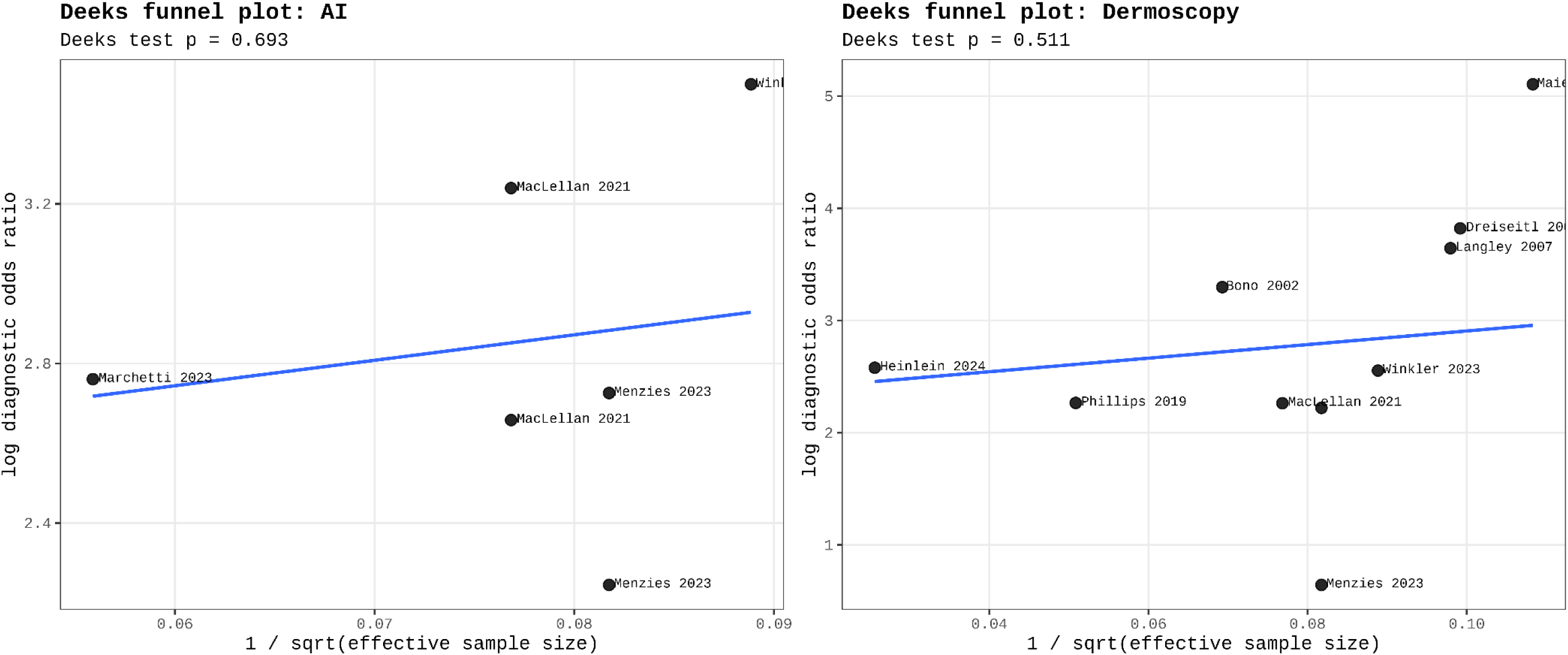
Deeks funnel plots for publication bias. Deeks funnel plots were used to assess potential publication bias and small-study effects in the AI and dermoscopy groups. No significant publication bias was detected in either the AI group (P = 0.693) or the dermoscopy group (P = 0.511).

## 4. Discussion

### 4.1 Principal findings and clinical parity

This systematic review and meta-analysis provides a comprehensive evaluation of the diagnostic efficacy of standalone artificial intelligence (AI), conventional dermoscopy, and AI-assisted clinicians (“Doctor AI”) in the malignancy risk stratification of pigmented skin lesions. Our primary findings indicate that in prospective, real-world clinical settings, the overall diagnostic performance of autonomous AI systems is broadly comparable to that of standard dermoscopy, as evidenced by the significant overlap in their Summary Receiver Operating Characteristic (SROC) curves. Although algorithmic assessment achieved a marginally higher pooled specificity (0.859 vs. 0.793), it was offset by a lower pooled sensitivity (0.757 vs. 0.773). These overlapping performance bounds suggest that, as an independent clinical decision tool, current machine learning models do not yet offer a definitive advantage over the established standard of care.

### 4.2 The “performance gap” between retrospective and prospective evidence

In recent years, deep learning systems, particularly Convolutional Neural Networks (CNNs), have demonstrated transformative potential in dermatological image recognition[30][31]. Numerous retrospective studies utilizing historical image databases have reported that AI can achieve classification accuracies for melanoma that match or even exceed those of experienced board-certified dermatologists[32][33]. However, the present analysis corroborates a growing “translation gap”: when these models are transitioned from highly structured experimental datasets to the stochastic nature of prospective clinical practice, their perceived superiority often diminishes[34]. The high heterogeneity of lesion morphology in the real world, non-standardized lighting and imaging equipment, and the interference of complex, rare pathologies are primary drivers of this performance attrition when translating from “bench to bedside”[35][37].

### 4.3 Threshold effects and algorithmic heterogeneity

A pivotal discovery of this study is the fundamental difference in the drivers of heterogeneity between the two diagnostic modalities. In the dermoscopy cohort, the correlation coefficients of logit sensitivity and false-positive rates suggest that variance is primarily driven by non-threshold factors, such as clinician expertise, training background, and diverse patient demographics. Conversely, the standalone AI group exhibited a perfect positive correlation (r = 1.00), indicating that the observed heterogeneity is almost exclusively attributable to “threshold effects.” This suggests that the performance variance across different AI systems reflects developers’ distinct calibration strategies for operating cut-offs—prioritizing high sensitivity to minimize missed diagnoses versus high specificity to reduce unnecessary biopsies—rather than fundamental disparities in the intrinsic discriminatory capacity of the neural networks themselves. Consequently, there is an urgent need for precise calibration of AI probability outputs and the establishment of standardized clinical operating thresholds[38].

### 4.4 Dataset bias and medical equity

The limitations of AI performance must also be viewed through the lens of dataset representational bias. Current mainstream models rely heavily on a few large-scale, open-source repositories, such as the HAM10000 and ISIC archives[39][41]. Evidence suggests that dark skin tones (Fitzpatrick types IV–VI) are significantly underrepresented in these datasets, leading to a marked decrease in generalizability and a risk of algorithmic bias when applied to diverse global populations[42][43]. Furthermore, isolated image-based inputs deprive the models of critical clinical metadata—such as patient history, rate of lesion evolution, and personal or family history of skin cancer—which are essential for definitive diagnosis[44]. The absence of this clinical context likely imposes a “ceiling effect” on the diagnostic accuracy of standalone image-analysis AI.

### 4.5 Synergistic diagnosis and the promise of “doctor AI”

Given these constraints, the narrative of AI as an autonomous replacement for human expertise should shift toward one of synergy. As advocated in this study, AI is currently best deployed as a synergistic decision aid. In our analysis, the single diagnostic arm evaluating this “human-computer collaboration” demonstrated superior metrics (sensitivity 1.000, specificity 0.837). While this finding necessitates cautious interpretation due to the limited evidence base, it underscores the potential of AI to bridge the experience gap between general practitioners and specialist dermatologists. By providing objective second opinions or flagging high-risk morphological features, AI can optimize diagnostic decision-making, maintaining high sensitivity while improving specificity to reduce the economic and physical burden of excising benign mimickers[33][37].

### 4.6 Strengths and limitations

This study strictly focused on prospective clinical trials, effectively filtering out the “laboratory bias” inherent in pure algorithm development studies. However, several limitations remain. First, most included studies were judged as high risk for patient selection bias, as they enrolled patients already identified with suspicious lesions rather than unselected screening populations. Second, the diversity of AI architectures and the relative scarcity of head-to-head “Doctor AI” trials limit our ability to draw definitive conclusions about the optimal integration strategy.

## 5 Conclusion

In conclusion, this systematic review and meta-analysis indicates that standalone AI demonstrates diagnostic performance broadly comparable to conventional dermoscopy for the malignancy risk stratification of pigmented skin lesions; however, current evidence does not substantiate the consistent superiority of autonomous AI. The marked variability across AI systems underscores that algorithmic efficacy remains highly context-dependent. While AI-assisted clinical assessment appears highly promising, the supporting evidence base is still nascent. At present, AI should be conceptualized not as a replacement for dermatological expertise, but as a supplementary tool to optimize diagnostic decision-making. Before AI can be seamlessly integrated into routine melanoma diagnostic pathways, future research must prioritize prospective multicenter designs, diverse patient cohorts, transparent algorithmic reporting, standardized operating thresholds, and clinically meaningful outcomes.

## Data Availability Statement

All data used in this systematic review and meta-analysis were extracted from previously published studies. The extraction form and all calculated diagnostic accuracy metrics are provided as Supporting Information. The original studies are cited in the reference list. No new primary data were collected in this study.

## Author Contributions

Hailing Tang: Conceptualization, Data curation, Formal analysis, Investigation, Methodology, Writing – original draft, Writing – review & editing.

Yaxi Zhu: Data curation, Investigation, Validation, Writing – review & editing.

Ming Diao: Conceptualization, Methodology, Supervision, Validation, Writing – review & editing.

## Ethics Statement

This systematic review and meta-analysis did not involve the collection of primary data from human participants or animals. All data were extracted from published studies. Therefore, ethical approval was not required. The study was registered with PROSPERO (CRD420261389834).

## Supporting Information

S1 File. PRISMA 2020 checklist.

S2 File. Full search strategies for all databases.

S3 File. Data extraction form and reconstructed 2×2 diagnostic tables for each included study arm.

